# Comparing the Demographic and Health Survey’s timing-based measure of unintended pregnancy to the London Measure of Unplanned Pregnancy in Bangladesh

**DOI:** 10.1101/2023.05.17.23290108

**Authors:** Md Nuruzzaman Khan, Shimlin Jahan Khanam, Melissa L. Harris

**Author notes:** **Corresponding author** Md Nuruzzaman Khan, Ph.D., Assistant Professor Department of Population Science, Jatiya Kabi Kazi Nazrul Islam University, Trishal, Mymensingh, Bangladesh.

## Abstract

**Background:** Demographic and Health Survey’s timing-based measure is commonly used in Low and Middle Income Countries to estimate unintended pregnancy despite its limitations, including ambivalent responses and failure to consider the partner’s intention, while LMUP, which can address these limitations, is not widely used in LMICs and is yet to be administered in Bangladesh. This study compared unintended pregnancy rates measured by the DHS timing-based measure and LMUP, as well as explored the extent of discordance between the measures and their determinants.

**Methods:** A cross-sectional survey was conducted in four districts of Bangladesh using two-stage stratified random sampling. Data was collected from 1,200 postnatal women. The study focused on discordance in reporting pregnancy intention between DHS timing-based measure and LMUP. Multivariate logistic regression models were used to identify predictors of discordant responses in reporting pregnancy intention.

**Results:** The prevalence of unintended pregnancy was found to be 24.3% through the DHS timing-based measure and 31.0% through the LMUP. Discordance in responses to pregnancy intention in the two measures was around 28%. Key predictors of discordance included older age, female last child, more than two children, poorer wealth quintile, and rural residence. Conclusions: Prevalence of unintended pregnancy in Bangladesh and other LMICs, measured by DHS timing-based measure, may grossly underestimated. This suggests that the negative effects of unintended pregnancy are even more significant than currently believed, further highlighting the need to strengthen the family planning program in Bangladesh.

## Background

Globally, almost half (47%) of all pregnancies, equivalent to approximately 121 million, are unintended, with low- and middle-income countries (LMICs) experiencing the majority of these cases due to lower usage of long-acting modern contraception and higher unmet need for contraception (1, 2). In particular, unintended pregnancy remains a key concern in Bangladesh, with over 47% of pregnancies at conception and around 23% of live births reported as unintended (3). Unintended pregnancies have significant negative consequences for maternal and child health, especially in LMICs, such as Bangladesh, where safe abortion services are restricted by law (1, 4). Approximately two-thirds of total unintended pregnancies in LMICs and Bangladesh end by induced abortions, contributing to around 13% of the total maternal mortality and costing billions of dollars in the treatment of abortion-related complications (1, 5). Unintended pregnancies resulting in live births can lead to delayed or inaccessible intra-partum and post-partum care, increasing the risk of several maternal and child health outcomes, such as stillbirths, preterm birth, low birth weight, child undernutrition, neonatal, and maternal mortality (3-7). Studies conducted in LMICs and Bangladesh have also found intergenerational effects of unintended pregnancy, such as higher rates of school dropout (8-10). Improving access to modern contraception and comprehensive sexuality education is crucial to reducing unintended pregnancies in Bangladesh and other LMICs (3).

The accuracy of current estimates of unintended pregnancy is a global concern, especially in LMICs where prospective data on reproductive events (e.g., conception, birth) are scarce, and data of unintended pregnancy usually come from the nationally representative Demographic and Health Survey (5). The survey collects timing-based data on pregnancy intention using two-time dependent questions which have several limitations. Specific limitations to this timing-based data include recall bias, which arises from difficulties in accurately remembering pregnancy intentions at the time of conception. Additionally, it can lead to a higher chance of ambivalent responses, particularly when the child’s sex is opposite to parental desire or when pregnancy intentions change after having a child (5). The major reasons for such limitations in data include: (i) they only apply to women who have given birth (i.e., excluding women who have had induced abortions), (ii) they do not include partners’ intentions, despite often being the dominant group in deciding pregnancy timing and number, and (iii) they do not consider contraceptive use/non-use status at the time of conception (11, 12). These factors can further result in ambivalence, denial, and confusion about pregnancy and misreporting of partner’s intentions, as a significant portion of reported unintended pregnancies in LMICs is wanted from the husband’s perspective (5, 11, 12).

To comprehensively measure pregnancy intention, it is necessary to consider multiple dimensions, such as contraception use, feelings about pregnancy at the time of conception and after giving birth, and agreement with the partner (12). The London Measure of Unplanned Pregnancy (LMUP) can cover these issues and can overcome the limitations of the DHS timing-based measure(13). However, the use of LMUP in LMICs is still limited and has not been included in national-level survey (11-14). Furthermore, there has been no previous investigation into the discordance between the DHS timing-based measure and LMUP or the factors contributing to this discordance. The aim of this study is to estimate the prevalence of unintended pregnancy in Bangladesh using two measures: DHS timing-based estimate and LMUP. The study also seeks to determine the degree of discordance between the prevalence estimates obtained from these two measures and identify the factors that contribute to such discordance.

## Methods

### Study settings and data

This study employed a cross-sectional survey design conducted in four randomly selected districts (Mymensingh, Pabna, Kurigram, Satkhira) using a two-stage stratified random sampling method. In the first stage, a total of 20 hospitals were selected. These comprised district level hospitals, one upazila health complex, one union health complex, and two private hospitals. Data were collected from 1200 women who met the inclusion criteria: having recently delivered (either by caesarean section or vaginal delivery) and were visiting the selected hospitals for post-natal healthcare services. The sample size required for this study was determined to be 378 using a sample determination formula (15). Data were collected through a face-to-face interview by using a pre-developed and pre-tested structured questionnaire. The questionnaire contained questions pertaining to both the DHS timing-based measure of unintended pregnancy and LMUP (16). In addition, other relevant questions from the Bangladesh Demographic and Health Survey, which is a part of the worldwide recognized Demographic and Health Survey program of the USA, were included. The questionnaire was initially developed and field-tested, with subsequent refinements made based on feedback received. The final version of the questionnaire and the data collection methodology were approved by the Institutional Ethical Board of the Institution of Biological Science at the University of Rajshahi, Bangladesh.

### Outcome measure

Discordant responses in pregnancy intention (yes, no) in DHS timing-based measure and LMUP was our outcome of interest. The variable was generated by comparing pregnancy intention estimates produced through the DHS timing-based measure and the LMUP. In the DHS-timing based measure, eligible women (those who had at least one live birth within five years of the survey) were asked two questions. The first question inquired whether they intended to become pregnant at the time of conception, with response options of “yes” or “no.” If the answer was “no,” a follow-up second question was asked to determine whether the unwantedness was for a short period (response option “later”) or permanent (response option “not at all”). We categorized these responses into intended pregnancy (positive response to the first question) and unintended pregnancy (negative response to the first question and “later” or “not at all” in the second question) (see Supplementary Table 1). On the other hand, total of six questions were asked to record pregnancy intention through LMUP. Based on the response to six questions and corresponding scores, the LMUP produces estimates for unplanned pregnancy (score: 0-3), ambivalence (score: 4-9) and planned pregnancy (score: 10-12) (Supplementary table 2). We considered the responses to be concordant in pregnancy intention if women consistently reported a similar type of pregnancy intention in both measures, such as intended pregnancy and planned pregnancy or unintended pregnancy or unplanned pregnancy. Any mismatch between these two measures, such as intended pregnancy and ambivalence or intended pregnancy and planed pregnancy, were considered discordant pregnancy intention responses.

### Explanatory variables

Explanatory variables were selected in two stages. Firstly, a list of potential explanatory variables were compiled by reviewing the relevant literature in LMICs, particularly Bangladesh. As this is the first study of its kind in LMICs, we considered similar research, such as studies investigating determinants of unintended pregnancy measured through the DHS timing-based measure or the LMUP (3, 5, 12-14, 17, 18). Next, we checked the availability of the selected variables in the survey and created a list of available variables. The selected variables included women’s age (categorized as ≤19 years, 20-34 years, and ≥35 years), women’s education (categorized as no formal education, primary education, secondary education, and higher education), women’s formal work engagement (yes or no), number of children ever born (categorized as 1-2 children and >2 children), partner’s education (categorized as no formal education, primary education, secondary education, and higher education), partner’s occupation (categorized as agriculture worker, physical worker, service, business, and others), sex of the last child (male or female), household wealth quintile (categorized as poorer, poorest, middle, richer, and richest), place of residence (urban or rural), and district of residence (Pabna, Mymensingh, Satkhira, and Kurigram).

### Statistical analysis

Descriptive statistics were used to depict the baseline characteristics of the study participants. To present the discordant responses in pregnancy intention between the DHS timing-based measure and the LMUP, we utilized cross-tabulation. Multivariate logistic regression model was used to explore the factors associated with pregnancy intention discordance. We estimated both crude and adjusted associations while ensuring that multicollinearity was addressed prior to running the model. The results were reported as odds ratios (ORs) along with their corresponding 95% confidence intervals (CIs). The statistical software package Stata (version 15.2) was used for all statistical analyses.

## Results

Data were collected from a total of 1200 respondents, and the background characteristics of the participants are summarized in Table 1. The majority of respondents were aged between 20-34 years (80.2%), had secondary level education (40.0%), and were not formally employed (69.0%). The majority of partners had a higher education level (36.17%) and reported business as their occupation (28.3%). The study population had an almost equal proportion of male and female children (49.0% and 51.0%, respectively). The majority of participants lived in rural areas (78.7%).

**Table 1:** Background characteristics of the respondents, N=1200.

| Characteristics | N (%) |
| --- | --- |
| **Mother's age at birth of the last child** |  |
| ≤19 years | 123 (10.3) |
| 20-34 years | 962 (80.2) |
| ≥35 years | 115 (9.58) |
| **Mothers' education** |  |
| No formal education | 115 (9.6) |
| Primary | 233 (19.4) |
| Secondary | 480 (40.0) |
| Higher | 372 (31.0) |
| **Mother's formal employment status** |  |
| Yes | 372 (31.0) |
| No | 828 (69.0) |
| **Partner's education status** |  |
| No formal education | 180 (15.0) |
| Primary | 211 (17.6) |
| Secondary | 375 (31.25) |
| Higher | 434 (36.17) |
| **Partner's occupation** |  |
| Agriculture worker | 183 (15.3) |
| Physical worker | 262 (21.8) |
| Services | 335 (27.9) |
| Business | 340 (28.3) |
| Other | 80 (6.7) |
| **Sex of the last child** |  |
| Male | 588 (49.0) |
| Female | 612 (51.0) |
| **Number of children ever born** |  |
| 1-2 children | 892 (74.3) |
| >2 children | 308 (26.7) |
| **Household wealth status** |  |
| Poorest | 238 (19.8) |
| Poorer | 240 (20.0) |
| Middle | 243 (20.3) |
| Richer | 237 (19.8) |
| Richest | 242 (20.2) |
| **Place of residence** |  |
| Urban | 256 (21.3) |
| Rural | 944 (78.7) |
| District |  |
| Pabna | 306 (25.5) |
| Mymensingh | 296 (24.7) |
| Satkhira | 310 (25.8) |
| Kurigram | 288 (24.0) |

The prevalence of unintended pregnancy, measured using the DHS timing-based measure and the LMUP, are presented in Supplementary file 1 and 2, respectively. A higher prevalence of unplanned pregnancy was found using the LMUP (31.0%) compared to the DHS timing-based measure (24.3%). Nearly 47.5% of women intended to become pregnant at the time of conception, and almost 53% reported that their pregnancy occurred at the right time. Additionally, approximately 29% of women reported that they had never discussed pregnancy with their partner, and around 43% reported that they did not take any health actions (such as, took folic acid, sought medical/health advice) before becoming pregnant (Figure 1).

**Figure 1:**
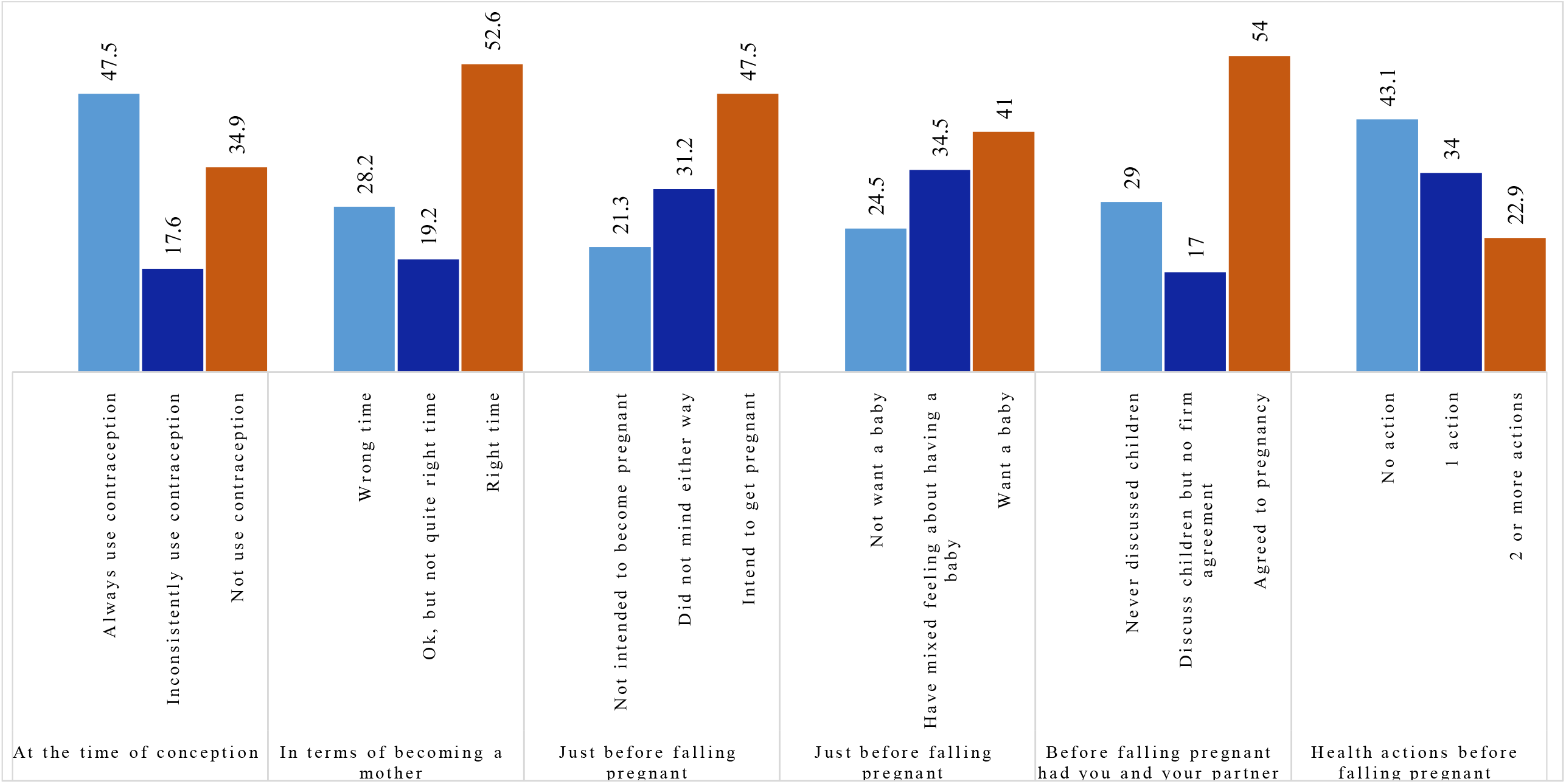
Women’s response to different questions on London Measure of Unintended Pregnancy

A total of 27.8% of the responses to pregnancy intention were discordant between the DHS timing-based measure and the LMUP (see Table 2).

**Table 2:** Comparison of pregnancy intention measured by Demographic and Health Survey’s timing-based measure and London Measure of Unplanned Pregnancy.

| LMUP<br>classifications* | DHS timing-based measure |  | Total |
| --- | --- | --- | --- |
|  | Unintended | Intended |  |
| Unplanned | 194 (CR) | 179 (DR) | 373 (31.0) |
| Ambivalent | 30 (DR) | 58 (DR) | 78 (6.5) |
| Planned | 68 (DR) | 681 (CR) | 749 (62.4) |
| Total | 292 (24.3) | 908 (75.7) |  |
| **Comparison in DHS timing-based measure and LUMP** |  |  |  |
| Concordant responses |  | 865 (72.2) |  |
| Discordant responses |  | 335 (27.8) |  |

**Note:** \*Classified based on scores presented in Supplementary table 2. Score 0-3: unplanned pregnancy; score 4-9: ambivalent; score 10-12: planned pregnancy. CR: Concordant response in both measures, DR: Discordant responses between two measures

Table 3 presents the factors associated with discordant responses in pregnancy intention, as determined by both crude and adjusted ORs. Increasing age of the respondent, female sex of the last child, having more than two children, being in the poorest wealth quintile, and residing in Kurigram district were associated with higher likelihood of discordant responses in the crude estimate. These findings remained consistent after adjusting for potential confounding factors in the multivariate model.

**Table 3:** Factors Associated with Discordance Responses in Pregnancy Intention Measured through Demography and Health Survey’s timing-based measure and London Measure of Unplanned Pregnancy Using Multivariate Logistic Regression Model.

|  | **Discordance response,**<br>**Crude odds ratio (95%**<br>**CI)** | **Discordance response,**<br>**Adjusted odds ratio**<br>**(95% CI)** |
| --- | --- | --- |
| **Mother's age at birth of the last child** |  |  |
| ≤19 years | 1.00 | 1.00 |
| 20-34 years | 1.28** (1.10 -1.49) | 1.32** (1.06 -1.53) |
| ≥35 years | 1.45* (1.08 -1.92) | 1.38* (1.06 -1.88) |
| **Mothers' education** |  |  |
| No formal education | 1.00 | 1.00 |
| Primary | 0.78 (0.63-1.26) | 0.82 (0.67 - 1.42) |
| Secondary | 0.94* (0.82 - 0.99) | 0.96* (0.80 - 0.95) |
| Higher | 0.82** (0.68-0.96) | 0.84* (0.63 - 0.95) |
| **Mother's formal employment status** |  |  |
| Yes | 1.00 | 1.00 |
| No | 1.14 (0.96-1.32) | 1.15** (1.01 - 1.38) |
| **Partner's education status** |  |  |
| No formal education | 1.00 | 1.00 |
| Primary | 0.93 (0.83 - 1.03) | 0.96 (0.85 - 1.07) |
| Secondary | 0.91 (0.81 - 1.04) | 0.95 (0.85 - 1.11) |
| Higher | 0.86 (0.78 - 0.94) | 0.89** (0.79 - 0.98) |
| **Partner's occupation** |  |  |
| Physical worker | 1.00 | 1.00 |
| Agriculture worker | 1.23 (0.93-1.53) | 1.26 (0.95-1.56) |
| Services | 1.13 (0.80 -1.46) | 1.18 (0.85 -1.48) |
| Business | 1.15 (0.84 – 1.46) | 1.18 (0.82 – 1.48) |
| Other | 1.06 (0.92 – 1.20) | 1.11 (0.88 – 1.26) |
| **Sex of the last child** |  |  |
| Male | 1.00 | 1.00 |
| Female | 1.38** (1.08 – 1.72) | 1.34** (1.06 – 1.67) |
| **Number of children ever born** |  |  |
| 1-2 children | 1.00 | 1.00 |
| >2 children | 1.24** (1.09 – 1.39) | 1.16* (1.07 – 1.40) |
| **Household wealth status** |  |  |
| Poorest | 1.39** (1.05 – 1.78) | 1.32** (1.06 – 1.82) |
| Poorer | 1.31** (1.02 – 1.64) | 1.34** (1.06 – 1.76) |
| Middle | 1.00 | 1.00 |
| Richer | 0.95 (0.86 - 1.04) | 0.98 (0.87 - 1.08) |
| Richest | 0.91* (0.87 – 0.95) | 0.94 (0.87 – 1.01) |
| **Place of residence** |  |  |
| Urban | 1.00 | 1.00 |
| Rural | 1.26 (1.04 – 1.48) | 1.30** (1.03 – 1.51) |
| **Districts** |  |  |
| Pabna | 1.00 | 1.00 |
| Mymensingh | 0.98 (0.92 -1.06) | 0.99 (0.93 -1.05) |
| Satkhira | 1.00 (0.99-1.03) | 1.03 (0.98-1.05) |
| Kurigram | 1.06* (1.01 - 1.10) | 1.08** (1.02 - 1.17) |

Mothers aged 20-34 and ≥35 years had 32% (95% CI, 1.06-1.53) and 38% (95% CI, 1.06-1.88) increased odds of having a discordant responses in pregnancy intention, respectively, compared to those aged ≤19 years. On the other hand, respondents with secondary (OR, 0.96, 95% CI, 0.80 - 0.95) and higher (OR, 0. 84, 95% CI, 0.63 - 0.95) education had decreased odds of having a discordant responses compared to those with no formal education or primary education. A 15- fold increase (ORs, 1.15, 95% CI, 1.01 - 1.38) in discordant responses was found among not formally employed mothers compared to employed mothers. Furthermore, mothers whose partners were higher educated (aOR, 0.89, 95% CI, 0.79 - 0.98) had lower odds of reporting discordant responses in pregnancy intention than mothers whose partners were not formally educated. Those having more than two children had a 16% (aOR, 1.16, 95% CI, 1.07-1.40) increased odds of having a discordance responses compared to those with 1-2 children. The poorest (aOR, 1.32, 95% CI, 1.06 – 1.82) and poorer (aOR, 1.34, 95% CI, 1.06 – 1.76) mothers were more likely to report discordance responses in pregnancy intention than middle wealth quintile mothers. Rural mothers were 30% (aOR, 1.30, 95%CI, 1.03 – 1.51) more likely to report discordance responses in pregnancy intention than urban mothers. Lastly, mothers in Kurigram district had an 8% (aOR, 1.08, 95% CI, 1.02-1.17) increased odds of discordant responses in pregnancy intention compared to those in Pabna district.

## Discussion

This study investigated unintended pregnancy prevalence using two measures - DHS timing-based and LMUP - and explored the degree of discordance between these measures, as well as the determinants of discordant responses. The study reveals that the prevalence of unplanned pregnancy reported through the LMUP (31.0%) was higher than through the DHS timing-based measure (24.3%). Additionally, approximately 28% of women provided discordant responses to pregnancy intention between the two measures. Significant predictors of discordant responses were increasing age, female sex of the last child, women having more than two children, being in the poorest wealth quintile, and residing in Kurigram district. These findings suggest that the actual occurrence of unintended pregnancy in Bangladesh and other LMICs may be higher than currently estimated through the DHS timing-based measure. Therefore, the burden on maternal and child health in LMICs related to unintended pregnancy may be even more significant than what is currently believed.

The reported prevalence of unintended pregnancy measured through the DHS-timing based measure in this study is consistent with reported national estimates in Bangladesh, where the same approach is adopted, as with other LMICs (1, 3, 17, 18). However, the use of a more comprehensive unintended pregnancy measure, the LMUP, revealed a much higher prevalence of unplanned pregnancy, with almost 38% of all pregnancies either unplanned (31%) or ambivalent (7%). In particular, individual dimensions of the LMUP revealed a more complex picture of unintended pregnancy, with only 54% of total pregnancies occurring with mutual agreement between the husband and wife. Furthermore, 43% of women reported not taking any prior health actions before getting pregnant, indicating a lack of adequate counselling on family planning and pre-pregnancy health check-ups. We could not validate these findings because of a lack of relevant literature in Bangladesh and LMICs. However, these findings highlight the urgent need for more comprehensive family planning services that address social norms and misconceptions around pregnancy, as well as provide counselling on family planning and preconception care (11, 12). Failure to address these issues can lead to poor maternal and child health outcomes in Bangladesh and other LMICs.

Our findings highlight that mixed feelings or uncertainty about pregnancy intention were common among women in Bangladesh, with one in four women reporting discordance. Young women and those with two or more children were more likely to report discordance in pregnancy intention, possibly due to their desire to delay or limit childbirth, respectively (3, 19-21). Additionally, women whose last child was female were more likely to report discordance, reflecting the persistent preference for male children in Bangladesh and other LMICs (22). This preference is especially prevalent among women from lower wealth quintiles and rural areas, who rely heavily on agriculture and view male children as primary workers in agriculture and primary caregivers in old age. This is in addition to perceptions surrounding the continuation of the family lineage and name(22-26). However, we also found that education played an important role in reducing discordance in pregnancy intention, as educated parents were less likely to prefer a child of a particular sex and were more likely to have access to family planning and contraception. As such, they appeared to have greater control over pregnancy planning (26-28).

The higher prevalence of discordance towards pregnancy intention suggests that a significant proportion of women have conflicting feelings about their pregnancy. This situation can have detrimental effects on various aspects. For instance, women who have contradictory feelings about their pregnancy are more likely to delay or have reduced access to maternal healthcare services, as evidenced by previous studies (3, 6, 7, 29). Additionally, women with unintended pregnancies or pregnancy ambivalence are more prone to engaging in negative behaviours during pregnancy, such as smoking, and have a higher prevalence of depression and anxiety(30). These concerns often stem from worries about their career or education(4). Collectively, these factors contribute to an increased risk of adverse maternal and child health outcomes, including high rates of maternal and child mortality already observed in Bangladesh and other LMICs (5, 8). Therefore, there is an urgent need for comprehensive family planning services and contraception, with a focus on comparatively young, poor, and less educated women. This direction should be incorporated into existing family planning services along with strengthening of current services.

This study has several key strengths including a large sample size. We exceeded the required sample size and increased the statistical power of our findings. Additionally, using two measures of unintended pregnancy, the DHS timing-based measure and the LMUP, provides a comprehensive understanding of the phenomenon. The study’s rigorous sampling method and inclusion criteria also enhanced internal validity and generalizability to the study population. Despite this, some limitations exist. The cross-sectional study design precludes establishing causal relationships in the reported associations in this study. The study’s inclusion of only women who had recently delivered and were receiving post-natal healthcare services at selected hospitals limited generalizability to the broader population of women in Bangladesh, particularly women who do not access delivery healthcare services and post-natal care. Reliance on self-reported interview data may also introduce reporting bias, as some women may have provided socially desirable responses. Lastly, the study did not collect data on other potential factors, such as social norms about child sex preference, that could influence discrepancies between the two measures of unintended pregnancy. Despite these limitations, this study is the first in LMICs to compare the commonly used DHS timing-based measure of unintended pregnancy with the more advanced LMUP, revealing a higher discordance rate. This study findings underscores the potential underestimation of the true prevalence of unintended pregnancy in Bangladesh when measured using the DHS-timing based measure. Consequently, the reported adverse outcomes associated with unintended pregnancy may be even higher than currently reported. This underscores the urgent need for effective family planning interventions and the promotion of modern contraception methods to ensure that all pregnancies are desired and planned. Considering that other low- and middle-income countries (LMICs) also employ similar measurement approaches for unintended pregnancy as Bangladesh, the findings of this study also emphasize the necessity for further research to accurately assess the extent of unintended pregnancy in LMICs and implement appropriate interventions accordingly.

## Conclusion

The prevalence of unplanned pregnancy estimated through the LMUP was 31%, which was almost 7% higher than the prevalence estimated through the DHS timing-based measure of 24.3%. About 28% of women provided discordant responses in pregnancy intention, with a significantly higher likelihood among women who were comparatively older in age, those whose last child was female, those who already had more than two children, and those in the poorest wealth quintile. These findings suggest that the actual extent of unintended pregnancy in Bangladesh may be higher than the currently available estimates derived from the DHS timing-based measure. As a result, the negative effects of unintended pregnancy may be more significant than what is currently perceived. Comprehensive programs that ensure family planning and contraception are necessary to empower couples to plan their pregnancies and to ensure every pregnancy is intended and conceived at the right time. Further, a comprehensive national-level study employing advanced techniques, such as LMUP, should be conducted to accurately determine the prevalence of unintended pregnancy. The findings of such a study can provide valuable insights for the development of comprehensive programs aimed at reducing the occurrence of unintended pregnancy and its related adverse outcomes.

## Data Availability

All data produced in the present study are available upon reasonable request to the authors

## Ethics approval

Ethical approval for this study was obtained from Human Research Ethics Committee of the Jatiya Kabi Kazi Nazrul Islam University with the approval number JKKNIU2023-07. The research protocol, including the study design, data collection procedures, and informed consent process, was thoroughly reviewed and deemed ethically sound and compliant with all applicable regulations and guidelines.

## Acknowledgment

We acknowledge the support Department of Population Science at Jatiya Kabi Kazi Nazrul Islam University, Bangladesh where this study was conducted.

## Authors’ contribution

MNK designed this study, conducted data analysis. SJK write the first draft of this manuscript along with MNK. MLH critically reviewed this manuscript. All authors approve the final version of this manuscript.

