## Supplementary file for "Comparing the Demographic and Health Survey’s timing-based measure of unintended pregnancy to the London Measure of Unplanned Pregnancy in Bangladesh"

Supplementary table 1: Measuring pregnancy intention using London Measure of Unintended Pregnancy

| **London Measure of Unintended Pregnancy** | | |
| --- | --- | --- |
| **Question** | **Score** | **Percentage** |
| 1) In the month that I became pregnant (please tick the statement that most applies to you): |  |  |
| I/we were not using contraception | 2 | 34.9 |
| I/we were using contraception, but not on every occasion | 1 | 10.6 |
| I/we always used contraception, but knew that the method had failed (ie, broke, moved, came off, came out, not worked, etc) at least once | 1 | 7.0 |
| I/we always used contraception | 0 | 47.5 |
| 2) In terms of becoming a mother (first time or again), I feel that my pregnancy happened at the (please tick the statement that most applies to you): |  |  |
| right time | 2 | 52.6 |
| Ok, but not quite right time | 1 | 19.2 |
| Wrong time | 0 | 28.2 |
| 3) Just before I became pregnant (please tick the statement that most applies to you): |  |  |
| I intended to get pregnant | 2 | 47.5 |
| My intentions kept changing | 1 | 31.2 |
| I did not intend to get pregnant | 0 | 21.3 |
| 4) Just before I became pregnant (please tick the statement that most applies to you): |  |  |
| I wanted to have a baby | 2 | 41.0 |
| I had mixed feelings about having a baby | 1 | 34.5 |
| I did not want to have a baby | 0 | 24.5 |
| In the next question, we ask about your partner. This might be (or have been) your husband, a partner you live with, a boyfriend, or someone you’ve had sex with once or twice. |  |  |
| 5) Before I became pregnant (please tick the statement that most applies to you): |  |  |
| My partner and I had agreed that we would like me to be pregnant | 2 | 54.0 |
| My partner and I had discussed having children together, but hadn’t agreed for me to get pregnant | 1 | 17.0 |
| We never discussed having children together | 0 | 29.0 |
| 6) Before you became pregnant, did you do anything to improve your health in preparation for pregnancy? (Please tick all that apply): |  |  |
| took folic acid |  | 11.1 |
| Stopped or cut down smoking | 2= 2 actions | 0.00 |
| Stopped or cut down drinking alcohol | 1 = 1 action | 0.00 |
| Ate more healthily |  | 21.2 |
| Sought medical/health advice |  | 23.1 |
| Took some other action (please describe) |  | 1.5 |
| Or |  |  |
| I did not do any of the above before my pregnancy | 0 | 43.1 |

Supplementary table 1: Measuring pregnancy intention using Timing Based Measure of Unintended Pregnancy

| **Questions** | **Response** | **Percentage** |
| --- | --- | --- |
| When you got pregnant with [name of last child born within three years of survey date], did you want to get pregnant at that time? | Yes | 75.7 |
|  | No | 24.3 |
| Did you want to have a baby later on, or did you not want any (more) children? | Later | 13.1 |
|  | Did not want any more | 11.2 |
